# Perception Gaps in Anatomical Competence: A Multi-Stakeholder Assessment of Physical Therapy Graduate Preparedness and Clinical Capability

**DOI:** 10.64898/2026.03.06.26347754

**Authors:** Michael A. Pascoe

**Author notes:** Correspondence: 13121 E. 17th Avenue, Mailstop C244, Aurora, CO 80045, United States. Author Contributions: The study conception and design were performed by Michael A. Pascoe. Material preparation, data collection, and analysis were also conducted by Michael A. Pascoe. The first draft of the manuscript was written by Michael A. Pascoe, who also reviewed and approved the final manuscript.

## Abstract

**Purpose:** Human anatomy remains foundational to clinical practice, yet reduced instructional hours raise concerns about graduate competence and preparedness for patient care. Although trainees often report confidence, supervisors may perceive deficiencies, creating a gap between self-assessment and external evaluation. This study examined stakeholder perspectives on anatomical competence within physical therapy education to identify areas of discordance in perceived capability.

**Methods:** A cross-sectional web-based survey collected responses from 165 stakeholders associated with an entry-level Doctor of Physical Therapy program featuring a 16-week dissection curriculum. Participants rated four domains of anatomical competence using a 5-point ordinal scale. Group differences were analyzed with the Kruskal–Wallis test appropriate for ordinal data. This methodology ensured robust assessment of stakeholder perceptions and comparative analysis.

**Results:** Median ratings of preparedness and capability were 4 of 5 (quite prepared). Significant discordance emerged in three domains: recent graduates rated their foundational knowledge and ability to explain complex concepts to lay audiences higher than faculty or clinical instructors, whereas faculty expressed lower confidence in graduates’ ability to explain patient symptoms using anatomical principles. No significant differences were observed in the ability to describe structures by location, suggesting shared perceptions of basic anatomical understanding despite variation in applied reasoning.

**Conclusions:** Stakeholders generally viewed graduates as well prepared, yet disagreement persisted regarding clinical application of anatomical knowledge. Faculty skepticism about symptom explanation indicates that mastery of anatomy alone does not guarantee clinical reasoning. Curricular strategies emphasizing vertical integration and explicit connections between anatomical science and patient-centered reasoning may help bridge perception gaps and enhance professional competence.

## INTRODUCTION

### Anatomy education in the global healthcare landscape

Human anatomy has long been viewed as the indisputable foundation of health professional training and an essential pillar of safe medical practice (Toogood et al., 2017). For centuries, this discipline has served as the primary requirement for those entering surgical and highly interventional clinical fields (Older 2004). However, the last two decades have seen a near-universal reduction in the time and resources allocated to teaching gross anatomy across medical schools worldwide (Drake et al., 2009). This shift, frequently described as the “winds of change,” has been driven by an ever-expanding breadth of medical advances that necessarily compete for time among traditional curricular staples (Ahmed et al., 2010; Drake et al., 2002). As education theory has evolved away from didactic lectures and the perceived burden of isolated facts, instruction in human anatomy has often been a common casualty during the trimming of healthcare curricula (Toogood et al., 2017). While new laboratory methods and digital tools have been introduced to supplement this gap, their relative effectiveness in producing competent clinicians remains a point of intense debate among established clinical faculty and anatomists (Lazarus et al., 2012; Wilson et al., 2018).

### Perceptions of preparedness and the stakeholder gap

A major issue identified in current medical education research is the significant gap between how trainees and their supervisors perceive readiness for clinical work. While many trainees report feeling well-prepared upon graduation, their program directors and senior consultants often disagree (Fillmore et al., 2016). For example, in the United States, medical educators have reported that only 29% of residency directors felt their residents were adequately prepared in gross anatomy, even though the learners themselves expressed a high level of confidence in their knowledge (Fillmore et al., 2016). This discordance suggests that non-experts may either overestimate their skills or lack a full understanding of the depth of anatomical insight required for safe patient care (Savran et al., 2015). Similar trends have been observed internationally; studies in the United Kingdom found that nearly half of newly qualified doctors believed they were not taught enough anatomy (Fitzgerald et al., 2008), while foundation year doctors in England reported that their knowledge was least sufficient for surgical and orthopedic rotations (Lewis et al., 2016). In Saudi Arabia and Ireland, both students and specialists like radiologists emphasize that a lack of anatomical knowledge is a direct barrier to safe practice (Almizani et al., 2022; O’Keeffe et al., 2019). This global discordance between confidence and capability highlights a critical need to reassess how we measure preparedness (Farey et al., 2018; Nabil et al., 2014).

### Theoretical frameworks of anatomical competence

To address these challenges, researchers have proposed the framework of anatomical competence, which encompasses both a foundational knowledge of structures and the ability to effectively utilize that knowledge in the care of patients (Fillmore et al., 2016). The theory of knowledge encapsulation helps explain why experts and novices view anatomy differently (Savran et al., 2015). Experts often encapsulate basic science facts into clinical scripts or shortcuts, while students rely on detailed biomedical facts but struggle to link them to patient symptoms (Woods et al., 2007). This is further complicated by the transfer trap, where students find it difficult to move knowledge from the classroom to the clinic (Lazarus et al., 2012). Educational researchers suggest that this transfer of knowledge occurs only 10% to 30% of the time, often because basic science is not taught with enough clinical emphasis (Lazarus et al., 2012). When anatomy is taught as a standalone basic science devoid of clinical relevance, students may excel at memorization but fail in clinical reasoning (Savran et al., 2015). This concern is particularly high in musculoskeletal specialties, where a lack of anatomical insight is linked to an increased risk of procedural errors (Toogood et al., 2017).

### The landscape of physical therapy education

Despite these broad concerns, most existing data on anatomical preparedness focuses on medical students, and there is a distinct scarcity of information regarding physical therapy education (Pascoe & Rapport, 2022; Rompolski et al., 2023). Physical therapists must possess a deep and functional understanding of the musculoskeletal and nervous systems to competently evaluate and treat movement dysfunction across the lifespan (Giuriato et al., 2020). However, content determination in physical therapy programs is often left to a single anatomy instructor, who may be an academic anatomist without any clinical experience (Pascoe & Rapport, 2022). In the United States, it is estimated that approximately 50% of faculty teaching anatomy in entry-level physical therapist education programs do not hold a clinical degree (Mattingly & Barnes, 1994). This lack of clinical input can lead to curricula that over-emphasize low-level naming of structures while neglecting the functional application required in the workplace (Pascoe & Rapport, 2022).

In several regions, physical therapy students have already reported negative attitudes toward their anatomy education because they struggle to link theoretical classroom knowledge to real patient cases (Ojukwu et al., 2022). Studies in Nigeria found that students often viewed anatomy as merely the study of dead bodies rather than a foundation for clinical skills (Ojukwu et al., 2022). Similarly, nearly half of practicing physical therapists in Turkey reported that their anatomy training was clinically deficient, especially in terms of practical application (Duman et al., 2017). Similar patterns have been noted in other allied health fields, such as occupational therapy, where students often value anatomy more highly than those who have been practicing for several years (Legleiter 2023). These findings across multiple disciplines highlight a global need to reassess how well healthcare curricula are meeting the actual needs of practicing clinicians (Giuriato et al., 2020).

### Improving consistency through stakeholder assessment

There is a clear need to examine these perspectives across multiple stakeholder groups within a single curriculum to understand how well students are being prepared for the workforce (Pascoe & Rapport, 2022). Just as clinical practice guidelines aim to reduce variations in care through standardization, educational research methods can be employed to increase consistency among courses through consensus determination (Pascoe & Rapport, 2022). Previous work utilizing the Lawshe method (Lawshe, 1975) has begun to identify which anatomical learning objectives are truly essential for physical therapy (Pascoe & Rapport, 2022). However, identifying essential content is only the first step. There remains a critical need to understand if the physical therapy curriculum is actually producing graduates who feel capable of using their knowledge to explain patient symptoms or complex details to a layperson (Pascoe & Rapport, 2022). By looking at the opinions of those who teach (faculty), those who have just graduated, and those who supervise in the clinic (clinical instructors), we can identify exactly where the educational gaps are located (Pascoe & Rapport, 2022).

### Statement of purpose

The objective of this study was to examine ratings of preparedness and capability by multiple stakeholders of a professional, entry-level physical therapist education program. Specifically, this study sought to compare the perceptions of core faculty members, recent graduates, and external clinical instructors regarding the graduates’ anatomical knowledge base and their ability to apply that knowledge in a physical therapy clinical setting. The study focused on a traditional 16-week curriculum that utilized whole-body donor dissection, a method still considered the cornerstone of anatomy education by many clinicians (Rompolski et al., 2023). By utilizing a web-based survey to collect and analyze stakeholder views, this research aims to determine if a perception gap exists between the educational environment and the practical application of anatomical knowledge in the physical therapy workforce. These results are intended to provide evidence-based guidance for educators seeking to optimize the placement and clinical relevance of anatomy within professional programs.

## MATERIALS AND METHODS

### Study design

A cross-sectional survey study was conducted to examine stakeholder perceptions of graduate preparedness and the capability to apply anatomical knowledge in clinical practice. This specific design was selected to capture contemporaneous ratings from multiple stakeholder groups associated with a single entry-level Doctor of Physical Therapy (DPT) program. By focusing on a single institution, the investigation could more precisely examine how one specific 16-week curriculum translates into varied clinical outcomes (Pascoe & Rapport, 2022). This approach aligns with current educational research which suggests that the opinions of stakeholders at the interface of training and practice are essential for assessing anatomical competence (Fillmore et al., 2016).

### Setting and participants

The investigation took place within one accredited entry-level DPT program in the United States. Human anatomy in this curriculum was delivered as a foundational science over 16 weeks, with ten weeks in the first year and six weeks in the second year. This structure provided a model of vertical integration where knowledge is revisited as students advance through the program (Lazarus et al., 2012). A hallmark of this specific course was the inclusion of whole-body donor dissection. This traditional method is widely regarded by both clinicians and students as the cornerstone for gaining a necessary three-dimensional perspective of the human body (Older 2004; Rompolski et al., 2023).

Eligible participants included three distinct stakeholder groups: (1) core faculty members in the program (*n* = 28), (2) recent graduates of the program from the two most recent cohorts (*n* = 134), and (3) external clinical instructors who supervised students during their rotations (*n* = 247). All identified individuals (*N* = 409) were invited to participate. Because every eligible individual was contacted, this represented a census sampling approach rather than a sampled subset. This ensures that the results are likely representative of the entire associated community (Pascoe 2022). Participation was entirely voluntary.

### Survey instrument

Data were collected using a web-based survey developed through the Qualtrics platform (Seattle, WA, USA). The instrument was created based on established standards for educational research to ensure the accurate measurement of the intended constructs (Artino et al., 2014). The final tool included demographic items and four Likert-scale items rated on a five-point ordinal scale. For all items, a score of 1 indicated not at all prepared/capable, while a score of 5 represented extremely prepared/capable (Pascoe & Rapport, 2022).

The items were designed to move beyond simple factual recall and instead reflect the applied clinical use of anatomy (Savran 2015). They assessed: (1) adequacy of anatomical knowledge at graduation, (2) capability to explain patient symptoms using underlying anatomy, (3) ability to describe structures by location, and (4) capability to explain complex concepts to a layperson. Prior to distribution, the program faculty reviewed the instrument for clarity and to ensure it aligned with current curricular objectives.

### Data collection and ethics

The survey (see Appendix) was distributed electronically via institutional email lists over a three-month period from April to June 2018 (Pascoe & Rapport, 2022). Invitations included a brief description of the study and a secure link to the survey platform. To improve the response rate, two reminder emails were sent at spaced intervals. Responses were collected anonymously, and incomplete surveys were included in the final analysis provided the demographic and preparedness sections were finished. The study protocol was reviewed and considered exempt by the Colorado Multiple Institutional Review Board (Protocol #18–0514). No identifiable personal data were collected, and no incentives were provided to participants.

### Statistical analysis

Survey responses were treated as ordinal data. Descriptive statistics were calculated for each item, including medians and response distributions. Calculating means for Likert-type scales is often considered illegitimate because the intervals between categories cannot be presumed equal (Jamieson 2004). Therefore, the Kruskal–Wallis test was chosen to evaluate differences in ratings across the three independent stakeholder groups. Statistical significance was determined a priori at the *P* < 0.05 level. These nonparametric tests were conducted using SPSS (IBM Corp., Armonk, NY, USA) to ensure a sound treatment of uncertainties within the data set. This analytic approach allowed for a robust comparison of perception patterns across different levels of clinical experience.

## RESULTS

### Participant characteristics

Of the 409 stakeholders invited to participate, 165 completed the survey (response rate: 40%). Respondents represented all three stakeholder groups: faculty, recent graduates, and clinical instructors. Participants reported a range of professional experience and levels of post-professional training. Detailed demographic information is presented in Table 1.

**Table 1.** Summary of survey responses to the demographic information section, per group and overall. DPT = doctor of physical therapy; MPT = master’s in physical therapy; BSPT = bachelor of science in physical therapy; DSc = doctor of science; CE = clinical experience.

| Survey item | Faculty | Recent graduate | Clinical instructor | Overall |
| --- | --- | --- | --- | --- |
| Submissions included | 21 | 56 | 88 | 165 |
| Years of experience (mean) | 9.4 | 1.1 | 8.5 | 6.1 |
| Highest clinical degree | 10 DPT<br>3 MPT | 56 DPT | 69 DPT<br>15 MPT<br>4 BSPT | 135 DPT<br>18 MPT<br>4 BSPT |
| Specialist certification (N, % of group) | 12 (57%) | 3 (5%) | 30 (34%) | 45 (27%) |
| Content delivered in DPT curriculum | 6 clinical reasoning<br>12 foundational<br>12 patient management | -- | -- |  |
| Highest academic degree | 3 PhD only<br>3 PhD & DPT<br>2 DSc | -- | -- |  |
| Clinical instructor certifications | -- | -- | 3 advanced only<br>43 credentialed only<br>9 credent & advance<br>30 either<br>3 blank |  |
| Clinical experiences supervised | -- | -- | 12 CE I only<br>33 CE II only<br>32 CE III only<br>3 CE I & II<br>3 CE I, II, III<br>5 CE II & III |  |

### Overall preparedness and capability ratings

Across all respondents, the median rating for each of the four survey items was 4 on the 5-point scale. A rating of 4 corresponded to quite prepared or quite capable. Response distributions for each item are shown in Table 2.

**Table 2.** Summary of ratings on graduate preparedness and capability to apply anatomy knowledge, per group and overall. The number of submissions per group is given in the column header. Data are presented as median (range) with the percentage of responses that were a 4 or 5 (out of 5). *Denotes significance, less than ⊡ = 0.05. For all survey items, a five-point Likert scale was used with 1 = not at all prepared/capable; and 5 = extremely prepared/capable.

| Survey item | Faculty (n = 21) | Recent graduate (n = 56) | Clinical instructor (n = 88) | Overall (N = 165) | Kruskal–Wallis test *P* value |
| --- | --- | --- | --- | --- | --- |
| 1. How prepared are CU PT students in terms of adequate anatomical knowledge, either in clinic or in your class? | 4 (1-5)<br>86% | 4 (3-5)<br>95% | 4 (2-5)<br>82% | 4 (1-5)<br>88% | 0.048* |
| 2. How capable are CU PT students of explaining patient symptoms using underlying anatomy? | 4 (2-5)<br>62% | 4 (3-5)<br>73% | 4 (3-5)<br>73% | 4 (2-5)<br>71% | 0.008* |
| 3. How capable are CU PT students of describing anatomical structures by location? | 4 (3-5)<br>86% | 4 (3-5)<br>84% | 4 (3-5)<br>84% | 4 (3-5)<br>84% | 0.53 |
| 4. How | 4 (2-5) | 4 (3-5) | 4 (2-5) | 4 (2-5) | < 0.001* |
| capable are CU<br>PT students of<br>explaining<br>complex<br>anatomical<br>details to a<br>layperson? | 48% | 82% | 49% | 60% |  |

For the item assessing adequacy of anatomical knowledge at graduation, most respondents selected ratings of 4 or 5. Similar response patterns were observed for the ability to describe anatomical structures by location and for the capability to explain complex anatomical information to a layperson. Ratings for the ability to explain patient symptoms using underlying anatomy showed greater spread across response categories.

### Comparison across stakeholder groups

Median ratings by stakeholder group are reported in Table 2. Statistically significant differences were observed among groups for three of the four survey items. For perceived adequacy of anatomical knowledge, recent graduates reported higher ratings than other stakeholder groups (Kruskal–Wallis test, *P* = 0.048). A similar pattern was observed for the capability to explain complex anatomical details to a layperson (*P* < 0.001). For the item assessing the capability to explain patient symptoms using underlying anatomy, faculty reported lower ratings relative to other groups (*P* = 0.008). No statistically significant difference was found among stakeholder groups for the ability to describe anatomical structures by location (*P* = 0.53). Percentages of respondents selecting ratings of 4 or 5 for each stakeholder group and survey item are presented in Table 2 and illustrated in Figure 1.

**Figure 1.**
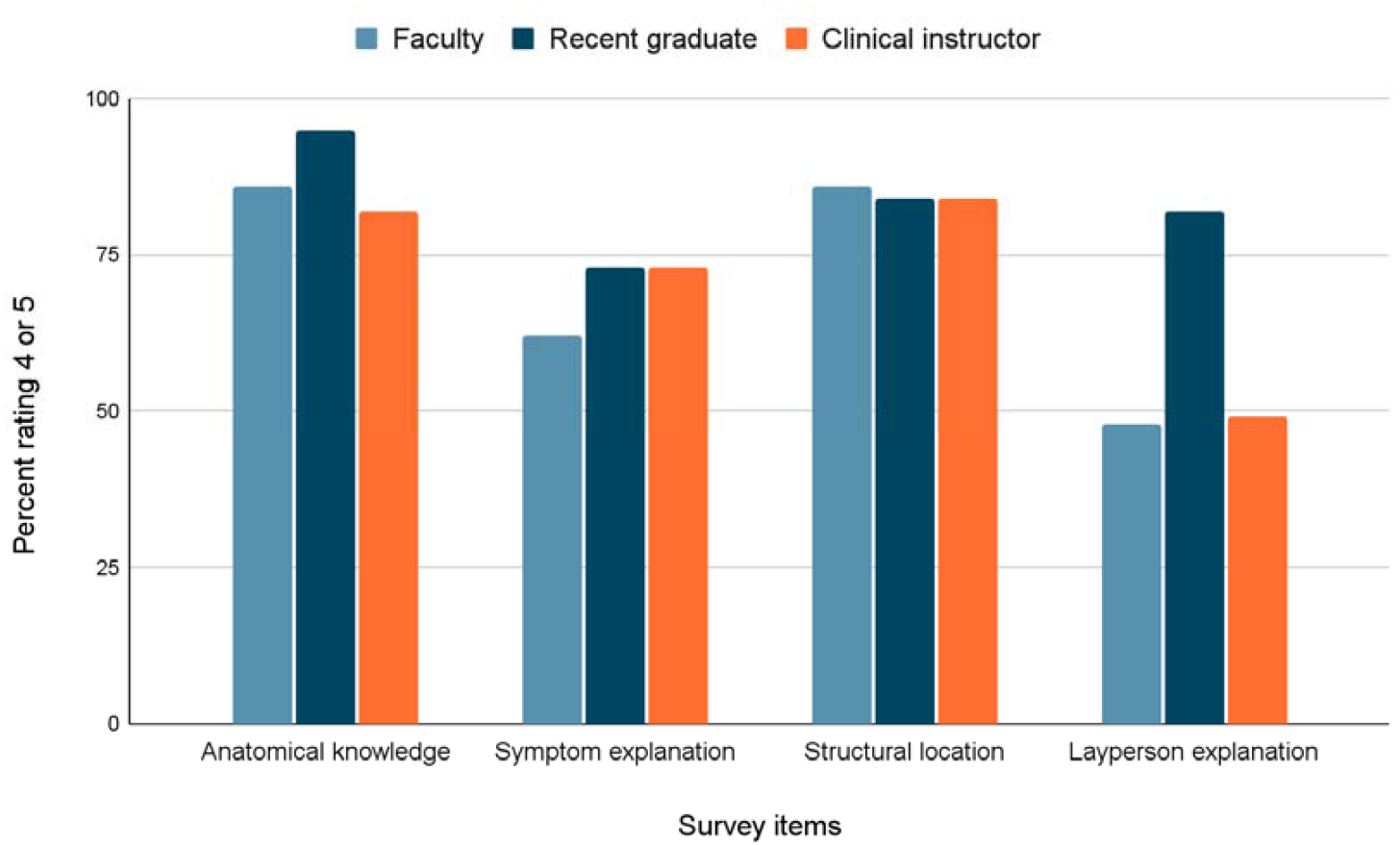
Stakeholder ratings reveal significant perception gaps between recent graduate confidence and clinical supervisor assessments of anatomical capability. The bars illustrate the percentage of respondents in each stakeholder group, faculty (*n* = 21), recent graduate (*n* = 56), and clinical instructor (*n* = 88), who assigned a rating of 4 or 5 on a 5-point Likert scale (where 1 = not at all prepared/capable and 5 = extremely prepared/capable). Data are compared across four distinct domains of anatomical competence: anatomical knowledge, symptom explanation, structural location, and layperson explanation. Statistically significant differences among the three independent groups (*P* < 0.05) were identified for all items except the description of structural location, as determined by the Kruskal–Wallis test.

## DISCUSSION

### Summary of primary findings

This investigation provides a multi-stakeholder assessment of graduate readiness to apply anatomical knowledge in a physical therapy setting. The overall results indicate a high level of perceived competence, with median ratings across all survey items reaching a 4 out of 5, which corresponds to being quite prepared or quite capable. This suggests that a traditional 16-week curriculum involving whole-body donor dissection is viewed by faculty, graduates, and clinical supervisors as effectively meeting the foundational needs of the profession. However, the data revealed significant areas of divergence among these groups. While the ability to describe anatomical structures by location was rated uniformly across all cohorts, significant differences were found in perceptions of foundational knowledge and the ability to explain clinical symptoms or complex details. These findings highlight a specific perception gap that must be addressed to optimize the transition from the classroom to the clinical environment.

### Stakeholder discordance in perceived preparedness

The highest ratings for anatomical preparedness and the capability to explain complex details to laypeople were provided by recent graduates. This finding aligns with established trends in medical education where learners frequently report higher levels of confidence than their supervisors (Fillmore et al., 2016; Nabil et al., 2014). In similar studies of medical residents, program directors consistently rated trainees as less prepared in gross anatomy than the residents rated themselves (Fillmore et al., 2016). This discordance may suggest that graduates are overestimating their readiness or, alternatively, that they lack an understanding of the depth of anatomical insight required for autonomous clinical decision-making (Savran et al., 2015).

Interestingly, these results contrast with some international data. For instance, nearly half of newly qualified doctors in the United Kingdom and physical therapy students in Nigeria have expressed dissatisfaction with the sufficiency of their anatomy training (Fitzgerald et al., 2008; Ojukwu et al., 2022). The high ratings observed in this study likely reflect the heavy musculoskeletal emphasis of the Doctor of Physical Therapy (DPT) curriculum, which directly supports the diagnostic and treatment needs of the profession (Giuriato et al., 2020; Rompolski et al., 2023).

### The theory of knowledge encapsulation

A critical finding of this study was that faculty members provided the lowest ratings regarding the graduates’ capability to explain patient symptoms using underlying anatomy. This specific gap is best explained by the theory of knowledge encapsulation (Savran et al., 2015; Woods et al., 2007). According to this framework, expert clinicians (such as faculty and experienced clinical instructors) integrate basic science facts into clinical scripts or cognitive shortcuts used during patient care (Savran et al., 2015). Novices, such as recent graduates, often possess elaborated biomedical knowledge but struggle with the transfer of that knowledge to real-world scenarios (Lazarus et al., 2012).

While the graduates in this study felt quite capable of reciting complex details, the experts who taught them were more skeptical of their ability to use that information for clinical reasoning. This confirms that mastering anatomy as a standalone science does not automatically translate into clinical competence (Fillmore et al., 2016). To bridge this gap, educators should consider more frequent use of clinically contextualized case studies and integrated laboratory experiences that require students to link structures directly to common movement dysfunctions (Pascoe & Rapport, 2022; Rompolski et al., 2023).

### Implications for physical therapy curricula

The results support the continued value of whole-body donor dissection as a cornerstone of physical therapy education. Respondents who participate in dissection often perceive it as essential for gaining a three-dimensional perspective that digital tools cannot yet fully replicate (Older 2004; Rompolski et al., 2023). However, given that faculty and supervisors remain concerned about clinical application, there is a clear need for vertical integration (Lazarus et al., 2012; Pascoe & Rapport, 2022).

Anatomy should not be a once and done course in the first year. Instead, programs should provide formal opportunities for anatomical review during clinical rotations or through advanced electives (Fillmore et al., 2016; Lewis et al., 2016). Stakeholders in this study, particularly clinical instructors, represent the front line of the profession. Their input is essential for identifying which content is truly essential and which areas are being over-taught at the expense of clinical reasoning (Pascoe & Rapport, 2022).

### Study limitations

Several limitations must be considered when interpreting these results. First, the investigation focused on a single entry-level physical therapist program, which limits the generalizability of the findings to other institutions with different laboratory formats or credit hours. Second, the cross-sectional nature of the study captures perceptions at one point in time and does not track the objective retention of knowledge over a long career. Third, all data were self-reported, which is subject to subjectivity and recall bias. Fourth, no a priori power analysis was performed, as the study was exploratory in nature. The sample size reflects all available respondents during the data collection period, and findings should be interpreted in the context of this descriptive design. Finally, the data were collected in 2018, and while the foundational nature of anatomy remains constant, the rapid evolution of educational technology during the COVID-19 pandemic may have altered more recent stakeholder perspectives (Rompolski et al., 2023).

## CONCLUSION

This study demonstrates that stakeholders generally perceive graduates of this physical therapist education program as well-prepared with the anatomical knowledge required for clinical practice. However, a significant perception gap exists between the confidence of the learners and the more conservative assessments of the faculty and clinical supervisors. These results emphasize that foundational knowledge alone is insufficient for professional competence. Future research should involve multi-center studies to determine if these perception patterns are consistent across different physical therapy curricula. Additionally, investigators should explore the correlation between these subjective stakeholder ratings and objective measures of clinical performance, such as licensure exam scores and patient care outcomes. By aligning the expectations of educators and clinicians, physical therapy programs can ensure that anatomy remains a functional and relevant pillar of clinical expertise.

## Supporting information

Appendix Full Survey Text

## Data Availability

All data produced in the present study are available upon reasonable request to the author.

## DECLARATIONS

### Ethics approval

The survey and data collection procedures were reviewed and considered exempt by the Colorado Multiple Institutional Review Board (Protocol #18-0514) in accordance with the ethical standards as laid down in the 1964 Declaration of Helsinki.

This manuscript is an original work and has not been published previously. The work is not under consideration in another journal. The author has no relevant financial or non-financial conflicts of interests to disclose. No funding was received for conducting this study. The author has no relevant financial or non-financial interests to disclose. Complete data sets are available upon request.

## Acknowledgments

The author would like to sincerely thank the faculty members, recent graduates, and clinical instructors who contributed their time and professional expertise by completing the survey instrument. A preliminary version of this study was presented in abstract form at the American Association for Anatomy (AAA) Annual Meeting, April 2022.

