## Appendix Full Survey Text for "Perception Gaps in Anatomical Competence: A Multi-Stakeholder Assessment of Physical Therapy Graduate Preparedness and Clinical Capability"

English

### Survey Information

### Survey Information

You are receiving this survey because you *1)* are CU PT Program Faculty, 2) are a recent Graduate of CU PT *or 3)* have worked with a CU PT student as their Clinical Instructor within the past 12 months. The purpose of this study is to examine faculty, recent graduate, & clinical instructor opinions of the anatomy curriculum in the University of Colorado Physical Therapy Program. This includes assessing how well the anatomy curriculum prepares students to succeed in other courses & in clinical education. Also of interest is your opinion of how essential certain course objectives are to providing clinical care to patients.

This survey is voluntary and will be anonymized after data collection. This survey will take approximately 10 minutes to compete.

This research is being conducted by individuals at the University of Colorado Anschutz Medical Campus, and is approved as an exempt study through the Colorado Multiple Institutes Review Board (COMIRB Protocol #18-0514). Please contact Mike Pascoe ( | 303.724.5978) with any questions or comments.

Begin the survey by selecting the button below.

### Section I: Demographic Variables

### Section I: Demographic Variables

What is your primary role?

- ○ CU PT Program Faculty
- ○ Recent CU PT Graduate (Class of 2017 or 2016)
- ○ Clinical Instructor

How many years of experience do you have in your primary role?

____________________

What is the highest PT degree you earned?

- ○ DPT
- ○ MPT
- ○ BSPT
- ○ __________ Other (please specify):

What academic degree(s) do you hold?

- ☐ PhD
- ☐ DPT
- ☐ EdD
- ☐ DSc
- ☐ MHS
- ☐ __________ Other (please specify):

Do you hold any of the following specialist certifications? Select any that apply.

- [x] Cardiovascular & Pulmonary
- [ ] Clinical Electrophysiology
- [ ] Geriatrics
- [ ] Hand Therapy
- [ ] Neurology
- [ ] Oncology
- [ ] Orthopaedics
- [ ] Manual Therapy Fellow
- [ ] Manual Therapy Certificate
- [ ] Pediatrics
- [ ] Sports
- [ ] Women's Health
- [ ] __________ Other (please specify):

What is the setting(s) of your current PT practice? Select all that apply.

- [ ] Acute Care/Inpatient Hospital Facility
- [ ] Ambulatory Care/Outpatient
- [ ] ECF/Nursing Home/SNF
- [ ] Federal/State/County Health
- [ ] Industrial/Occupational Health Facility
- [ ] Private Practice
- [ ] Rehabilitation/Sub-acute Rehabilitation
- [ ] School/Preschool Program
- [ ] Wellness/Prevention/Fitness Program
- [ ] Home Health/Hospice
- [ ] __________ Other (please specify):

Have you earned either of the following clinical instructor certifications?

- [ ] Credentialed CI
- [ ] Advanced Credentialed CI
- [ ] Neither

What clinical education experience(s) have you supervised in the past 12 months?

- [ ] CE I (May-June; 1st year student)
- [ ] CE II (Jan-March; 2nd year student)
- [ ] CE III (Aug-Dec; 3rd year student)

What type of content do you deliver in the CU PT curriculum? Select all that apply.

- [ ] Foundational Content (e.g., Clin Anat, Applied Exer Sci, EBP, Exam Eval, Exer Sci, FI, Human Growth Devel, Motor Control Learning, Move Sci, Neurosci, Prof Devel, Psycho Soc)
- [ ] Patient Management (e.g., HCD, Health & Wellness, Med Cond, MSK Cond, Neuromusc Cond)
- [ ] Clinical Reasoning (e.g., Clin Reasoning, Integrated Practice)
- [ ] ____________ Other (please specify):

### Section II: CU PT Student Preparedness

**Section II: CU PT Student Preparedness**

The questions in this section are related to how prepared CU PT students are for success related to their anatomical knowledge.

How prepared are CU PT students in terms of adequate anatomical knowledge, either in clinic or in your class?

| Not at all prepared | Slightly prepared | Moderately prepared | Quite prepared | Extremely prepared |
| --- | --- | --- | --- | --- |
| ○ | ○ | ○ | ○ | ○ |

How capable are CU PT students of explaining patient symptoms using underlying anatomy?

| Not at all capable | Slightly capable | Moderately capable | Quite capable | Extremely capable |
| --- | --- | --- | --- | --- |
| ○ | ○ | ○ | ○ | ○ |

How capable are CU PT students of describing anatomical structures by location?

| Not at all capable | Slightly capable | Moderately capable | Quite capable | Extremely capable |
| --- | --- | --- | --- | --- |
| ○ | ○ | ○ | ○ | ○ |

How capable are CU PT students of explaining complex anatomical details to a lay person?

| Not at all capable | Slightly capable | Moderately capable | Quite capable | Extremely capable |
| --- | --- | --- | --- | --- |
| ○ | ○ | ○ | ○ | ○ |

### Section III: Necessity of Anatomical Objectives

**Section III: Necessity of Anatomical Objectives**

Please select the option that best describes the importance of each anatomical objective below in your primary role (faculty / recent graduate / clinical instructor).

You may notice that foundational anatomical knowledge like anatomical directions and the names of muscles, bones, joints, nerves, and vessels are already considered essential and not included in the list.

Essential is defined as knowledge which you deem fundamental in providing clinical care to patients, communicating with other health care professionals, and/or keeping up with relevant literature.

### Cardiovascular System Objectives

| | Not Necessary | Useful but not Essential | Essential | Do Kn |
|---|---|---|---|---|
| 1. Explain fetal circulation | ○ | ○ | ○ | ○ |
| 2. Explain conduction system of the heart | ○ | ○ | ○ | ○ |
| 3. Explain the fetal development of the heart | ○ | ○ | ○ | ○ |
| 4. Identify cardiac auscultation sites | ○ | ○ | ○ | ○ |
| 5. Explain the venous pathways involved in each of the four primary portal hypertension scenarios (i.e., esophageal, rectal, lumbar, umbilical) | ○ | ○ | ○ | ○ |
| 6. List the contents of the four divisions of the mediastinum | ○ | ○ | ○ | ○ |

### Pulmonary System Objectives

| | Not Necessary | Useful but not Essential | Essential | Do Kn |
|---|---|---|---|---|

| | | | | |
|---|---|---|---|---|
| 7. Identify all 10 bronchopulmonary segments on both right and left lungs | ○ | ○ | ○ | ○ |
| 8. Identify the sinus cavities of the skull | ○ | ○ | ○ | ○ |
| 9. Describe the drainage of each sinus cavity into the nasal cavity | ○ | ○ | ○ | ○ |
| 10. Identify pulmonary auscultation sites | ○ | ○ | ○ | ○ |

### Neuromuscular & Nervous Systems Objectives

| | Not Necessary | Useful but not Essential | Essential | Do Kn |
|---|---|---|---|---|
| 11. Describe the seven functions an axon can have using functional component terminology (e.g., GSE, SVA) | ○ | ○ | ○ | ○ |
| 12. Draw and label the monosynaptic reflex pathway | ○ | ○ | ○ | ○ |
| 13. Draw and label all peripheral nerves arising from the brachial plexus (16 total) | ○ | ○ | ○ | ○ |
| 14. Name the branchial (pharyngeal) arches | ○ | ○ | ○ | ○ |
| 15. Name what structures derive from each branchial arch | ○ | ○ | ○ | ○ |
| 16. Describe the relative location of muscular nerve branch origin along the course of a peripheral nerve | ○ | ○ | ○ | ○ |
| 17. Explain the distribution of sympathetic innervation of the head | ○ | ○ | ○ | ○ |
| 18. List the structures that pass through the sciatic foramina | ○ | ○ | ○ | ○ |

### Musculoskeletal System Objectives

| | Not Necessary | Useful but not Essential | Essential | Do[illegible] Kn[illegible] |
|---|---|---|---|---|
| 19. Name every spinal segment present in a peripheral nerve | ○ | ○ | ○ | ○ |
| 20. Name every spinal segment that innervates a given skeletal muscle | ○ | ○ | ○ | ○ |
| 21. Use Hilton's Law to name all peripheral nerves that innervate a given joint | ○ | ○ | ○ | ○ |
| 22. Explain limb bud rotation of upper and lower extremities | ○ | ○ | ○ | ○ |
| 23. Identify the suboccipital muscles of the neck, their innervation, and function | ○ | ○ | ○ | ○ |
| 24. Name which of the four layers of muscles of the foot each intrinsic muscles belongs to | ○ | ○ | ○ | ○ |
| 25. Describe the retinacula of the knee | ○ | ○ | ○ | ○ |
| 26. Name the facial expressions produced by the muscles of facial expression | ○ | ○ | ○ | ○ |
| 27. Match layers of the anterior abdominal wall with their corresponding layer in the spermatic cord | ○ | ○ | ○ | ○ |
| 28. Explain the layers of the rectus sheath | ○ | ○ | ○ | ○ |
| 29. Explain the significance of the arcuate line | ○ | ○ | ○ | ○ |
| 30. Distinguish between the neurocranium and viscerocranium | ○ | ○ | ○ | ○ |
| 31. Name the bones that form the four walls of the orbit | ○ | ○ | ○ | ○ |
| 32. Explain how facet joint orientation limits motion in the three planes of movement, for the cervical, thoracic, and lumbar regions of the vertebral column | ○ | ○ | ○ | ○ |

### Lymphatic System Objectives

| | Not Necessary | Useful but not Essential | Essential | Do Kn |
|---|---|---|---|---|
| 33. Name all groups of lymph nodes in a given region | ○ | ○ | ○ | ○ |
| 34. Describe the territory each group of lymph nodes drain | ○ | ○ | ○ | ○ |
| 35. Describe the typical pattern of drainage for groups of lymph nodes | ○ | ○ | ○ | ○ |
| 36. Locate the parietal and visceral thoracic lymph nodes | ○ | ○ | ○ | ○ |

### Gastrointestinal System Objectives

| | Not Necessary | Useful but not Essential | Essential | Do Kn |
|---|---|---|---|---|
| 37. Name the three umbilical folds | ○ | ○ | ○ | ○ |
| 38. Name the contents of the three umbilical folds | ○ | ○ | ○ | ○ |
| 39. Distinguish between the arterial supply of the ileum and jejunum (e.g., vasa recta and arcades) | ○ | ○ | ○ | ○ |
| 40. Explain the importance of the pectinate line of the anus | ○ | ○ | ○ | ○ |

### Integumentary System Objectives

| | Not Necessary | Useful but not Essential | Essential | Do Kn |
|---|---|---|---|---|

| | | | | |
|---|---|---|---|---|
| 41. Describe anatomical components of the breast | ○ | ○ | ○ | ○ |
| 42. Explain the anatomy underlying lactation | ○ | ○ | ○ | ○ |
| 43. Summarize development of the breast (childhood to maturity) | ○ | ○ | ○ | ○ |

### Reproductive System Objectives

| | Not Necessary | Useful but not Essential | Essential | Do Kn |
|---|---|---|---|---|
| 44. Describe the curvature of the uterus (retro- and anteflexion, and retroversion) | ○ | ○ | ○ | ○ |
| 45. Define the boundaries of the superficial and deep perineal pouches of male and female | ○ | ○ | ○ | ○ |
| 46. Match the developmental homologues of the male and female external genitalia | ○ | ○ | ○ | ○ |

Are there any anatomical topics that stand out to you as **strengths of** the CU PT anatomy curriculum?

Are there any anatomical topics that stand out to you as **needing greater emphasis in** the CU PT anatomy curriculum?

Powered by Qualtrics
